# Association between insecticide-treated bed net use and Burkitt lymphoma incidence in children in sub-Saharan Africa: a systematic review and population-level analysis

**DOI:** 10.1101/2023.09.29.23296342

**Authors:** Nora Schmit, Jeevan Kaur, Elom K. Aglago

## Abstract

**Background:** Burkitt lymphoma (BL) is one of the most common childhood cancers in sub-Saharan Africa and aetiologically linked to malaria. However, evidence for an effect of malaria interventions on BL is limited. The aim of this study was to investigate the potential population-level association between large-scale rollout of insecticide-treated bednets (ITNs) in sub-Saharan Africa in the 2000s and BL incidence.

**Methods:** We conducted a systematic review in Embase, Global Health and MEDLINE to assemble all available data on BL incidence rates in children aged 0-15 years in malaria-endemic sub-Saharan African countries between Jan 1, 1990 and Feb 27, 2023. We calculated pooled estimates of BL incidence in sub-Saharan Africa for the time periods before and after ITN introduction. We used model estimates of sub-national ITN use to assess the association between average population ITN use and childhood BL incidence rates in a negative binomial regression model.

**Results:** We included 23 studies comprising 66 datapoints on BL incidence based on 5,226 cancer cases from locations with large-scale ITN use in 17 countries. BL rates were 44% (95% CI 12-64%) lower in the time period after ITN introduction compared to before. The pooled incidence rate of BL was 1.36 (95% CI 0.88-2.10) and 0.76 (95% CI 0.50-1.16) per 100,000 person-years before and after introduction of ITNs, respectively. After adjusting for potential confounders, a 1% increase in average ITN use in the population in the 10 years before BL data collection was associated with a 2% (95% CI 1-4%) reduction in BL incidence.

**Conclusion:** These findings suggest that large-scale rollout of ITNs in the 2000s was associated with a reduction in childhood BL burden in sub-Saharan Africa. Although published data may not be representative of all incidence rates across sub-Saharan Africa, our study highlights a potential additional benefit of malaria control programmes.

## Introduction

Burkitt lymphoma (BL) is an aggressive type of non-Hodgkin lymphoma and among the most common childhood cancers in sub-Saharan Africa (1). Infection with the ubiquitous Epstein-Barr virus (EBV) is an established risk factor for BL, and there is substantial epidemiological and biological evidence for an aetiological relationship between *Plasmodium falciparum* malaria and BL in sub-Saharan Africa (1, 2). Malaria still poses an important public health problem, with over 200 million clinical cases annually (3). Malaria parasite prevalence and the cumulative number of infections correlate with BL incidence on the population level (4, 5), and individual-level associations between malaria infection or anti-malarial antibody titres and BL risk have been reported (6).

This combined evidence raises the possibility that interventions against malaria might have downstream beneficial effects on reducing the burden of BL. In an uncontrolled malaria suppression trial in 1977-1982 in Tanzania, a substantial decline in BL incidence rates was observed during a mass chloroquine distribution campaign, starting before the trial years and subsequently rising to pre-trial levels (7). Two hospital-based case-control studies suggested vector control interventions could potentially decrease the risk of developing BL in African children [odds ratio (OR) 0.2 (95% CI 0.03-0.9) and 0.3 (95% CI 0.2–0.7) for children using mosquito nets or living in a household with insecticide use, respectively] (8, 9). In contrast, a recent large population-based case-control study reported an increased risk of BL in children having used a mosquito net the previous night (OR 2.4, 95% CI 1.8-3.2) and in children living in households exposed to indoor residual insecticide spraying in the past year (OR 2.3, 95% CI 1.8-3.1) (10). These mixed results may indicate a difficulty in accounting for residual confounding from malaria burden in the study of such a rare disease, and for the temporal relationship between intervention exposure and BL development (2, 10).

On the population level, the question of whether BL incidence has declined over time following scale-up of malaria interventions has not been investigated. The 2000s marked a period of unprecedented donor funding and the first time malaria interventions were rolled out in sub-Saharan Africa at a large scale (3). Mass campaigns focused on distributing insecticide-treated nets (ITNs) and artemisinin-based combination therapies (3). These control efforts reduced *P. falciparum* infection prevalence in sub-Saharan Africa by an estimated 50% between 2000 and 2015, with most of this decline attributable to ITNs (11).

In this study, we aimed to investigate the potential association between large-scale rollout of ITNs in the early 2000s and BL incidence in sub-Saharan Africa using an ecological study design. First, we conducted a systematic review to assemble all publicly available data on BL incidence rates in sub-Saharan African children since 1990. We calculated pooled estimates of BL incidence across malaria-endemic sub-Saharan African countries before and after large-scale ITN introduction and compared time trends in incidence in individual locations. Second, we used model estimates of sub-national ITN use (12) to investigate the association between average ITN use in the population and BL incidence in children.

## Methods

### Systematic review

The systematic review was conducted on cancer registry publications and in literature databases. The International Agency for Research on Cancer (IARC) collates data from population-based cancer registries globally; we assembled BL incidence data from publications identified through a search of

IARC-associated websites. We also searched EMBASE, Global Health and Medline databases up to 27^th^ February 2023 using the Ovid platform for all studies on BL incidence rates in sub-Saharan Africa published since 1^st^ January 1990 without language restrictions. Search terms included keywords for sub-Saharan African countries, BL and non-Hodgkin lymphoma and the outcome of interest (**Table S1**).

Epidemiological studies reporting the incidence rate of BL in children aged 0-15 years in malaria-endemic sub-Saharan African countries were included. Both population-based and hospital-based studies were considered for identification of cancer cases, but incidence rates had to apply to the general population. The outcome of interest was incident cancer cases diagnosed as BL in a defined population during a specified time period with a midpoint after 1990. Studies were eligible for inclusion if they reported the number of BL cases and either the person-time at risk or the crude incidence rate.

Articles were deduplicated and screened in the Covidence software. Screening was performed independently by two reviewers blinded to each other’s decision, and disagreements were resolved through discussion. One article reported BL incidence in 0-19 year while otherwise meeting inclusion criteria, but unpublished data in the 0-15 year group could not be obtained on request (4). Data was extracted independently by the two reviewers into a pre-validated Microsoft Office Excel template, with any differences resolved in a final assessment. Harmonised extracted data included study location, time, study design, data collection and ascertainment methods, age range, number of cases, person-years at risk, and incidence rate. Studies identified from the published literature were cross-checked against the dataset extracted from cancer registry publications for duplicates and overlap in data from the same registry. Their quality was scored based on three predefined criteria (data collection, case ascertainment, and calculation of person-time at risk).

This systematic review was registered in the PROSPERO register (reference CRD42023411961) and is reported according to PRISMA guidelines (13). Full methodological details can be found in the Supplementary Materials.

### Statistical analysis

Crude BL incidence rates in 0-15 year olds were converted to units of cases per 100,000 person-years. Due to the rarity of the cancer, BL incidence is usually reported for aggregate time periods. In the analysis, rates were assumed to apply to the calendar midpoint of the data collection period.

#### Time trends in BL incidence

To generate pooled incidence rates for sub-Saharan Africa in the period before and after large-scale ITN introduction, we estimated the year of ITN introduction for each location with BL data. The Malaria Atlas Project (MAP) generates comparable model estimates of yearly ITN use since 2000 at a fine spatial resolution (12). Based on information about the covered population in each study, we linked each BL datapoint to its corresponding first administrative level (admin1) unit. The urbanicity status of the population covered by the study was used to split ITN use estimates into urban and rural areas within each unit using a threshold density of 1500 people per square kilometre. We calculated the mean ITN use weighted by the population at risk in each location over time.

ITN use has generally increased since 2000, but temporal patterns vary between locations (**Figure S1**). To capture the beginning of large-scale population-wide ITN campaigns, we defined the year of ITN introduction as the first year with at least 5% usage and an increasing trend. Several southern African countries had nationwide BL data available but did not introduce ITNs at a large scale (mean ITN use below 1%) (14) (**Figure S1**). These countries employed other malaria interventions (e.g. artemisinin-based combination therapies and indoor residual spraying) (14); therefore, we considered their data separately.

BL incidence rates in each group (time period before and after ITN introduction, and in countries without large-scale ITN use) were analysed using a negative binomial regression model with person-time at risk as an offset and including location-level random intercepts. The pooled BL incidence rate with 95% confidence intervals was predicted for each group and for individual countries. Additionally, for the subset of locations with at least one BL datapoint before and after ITN introduction, we calculated the rate ratio for BL incidence after ITN introduction compared to before to assess trends in BL incidence for each location individually.

#### Population-level association between ITN use and BL incidence

To investigate the association between average ITN use in the population and BL incidence in children, we considered different time lags for population-level exposure to ITNs. The latency period between malaria exposure and BL onset is not fully understood: most children in highly endemic areas have their first exposure to both EBV and malaria in the first few years of life (15), while BL is most common in children aged 5-10 years (1). This observation led to the hypothesis that repeated malaria infections during childhood increase the risk of BL (5). Given this and the variable patterns in ITN scale-up (**Figure S1**), we hypothesised that ITN use in a particular year would be a poor predictor of BL incidence and instead used the mean population ITN use in the 10 years before BL data collection as measure of exposure. In sensitivity analyses, we also tested models with mean population ITN use in the 5 or 15 years before BL data collection, as well as concurrent ITN use. We assumed no exposure to ITN use in the population before the year 2000 (3).

We identified several *a priori* potential confounders for the population-level relationship between ITN use and BL incidence: admin1-specific baseline malaria prevalence in 2000 (before implementation of interventions) and concurrent HIV prevalence in adults, the country-specific human development index (HDI) and the urbanicity status of the population covered by the cancer registry or study (**Table 1**).

**Table 1.** Overview of model covariates and data sources. Summary statistics cover the 66 malaria-endemic locations in sub-Saharan Africa with data on Burkitt lymphoma incidence and large-scale insecticide-treated net (ITN) use. n = number of datapoints, IQR = interquartile range.

| Covariate | Description | Source | Before ITN introduction |  | After ITN introduction |  |
| --- | --- | --- | --- | --- | --- | --- |
|  |  |  | n (%) | Median [IQR] | n (%) | Median [IQR] |
| Average population ITN use over 10 years (%) | Mean of yearly location-specific ITN use in the population in the 10 years before (inclusive of) the midpoint year of the Burkitt lymphoma datapoint. ITN use represents the proportion of people who sleep under a net. | Malaria Atlas Project (12), *foresite* R package (16) | 35 (53) | 0.0 [0.0-0.8] | 31 (47) | 17.6 [11.0-29.0] |
| Baseline malaria parasite prevalence in 2000 (%) | Location-specific *P. falciparum* parasite prevalence in 2-10 year olds in 2000. The year 2000 is commonly considered as representing baseline endemicity levels before large-scale intervention use and marking the beginning of a historically unprecedented decline in malaria burden (51). Malaria prevalence correlates with Burkitt lymphoma incidence and is a key consideration in targeting malaria control programmes. | Malaria Atlas Project (52), *foresite* R package (16) | 35 (53) | 26.3 [22.2-48.7] | 31 (47) | 35.9 [19.5-50.8] |
| Concurrent HIV prevalence in 15-49 year olds (%) | Location-specific HIV prevalence in 15-49 year olds in the midpoint year of the Burkitt lymphoma datapoint as proxy for HIV prevalence in children. HIV infection is a strong risk factor for Burkitt lymphoma outside of sub-Saharan Africa, including in patients treated with antiretroviral therapy (53). HIV treatment programme scale-up also occurred in the 2000s and are supported by the same international funder as malaria (3, 54). | Dwyer-Lindgren et al 2019 (55) | 35 (53) | 7.8 [2.5-15.2] | 31 (47) | 5.3 [2.9-8.4] |
| Concurrent country Human Development Index (HDI) level | Country-specific human development index level in the midpoint year of the Burkitt lymphoma datapoint. The HDI is a summary measure of human development including life expectancy, education and income. HDI was | United Nations Human |  |  |  |  |
| *HDI < 0.45* | categorised into two levels at the median value in the dataset due to an observed non-monotonic relationship between HDI and BL incidence. HDI correlates with cancer patterns (56) and could be related to access to malaria interventions. | Development Reports (57) | 22 (33) | / | 9 (14) | / |
| *HDI ≥ 0.45* |  |  | 13 (20) | / | 22 (33) | / |
| Population urbanicity status | Urbanicity status of the population covered by the cancer registry or study. Most population-based cancer registries are located in cities and cover urban populations (31), while malaria is more common in rural areas (58). | Burkitt lymphoma data sources |  |  |  |  |
| *Urban* |  |  | 16 (24) | / | 18 (27) | / |
| *Both* |  |  | 14 (21) | / | 12 (18) | / |
| *Rural* |  |  | 5 (8) | / | 1 (2) | / |

We calculated crude and adjusted rate ratios for these predictors using a negative binomial model with person-time at risk as an offset and including geographical location as a random intercept to account for potential correlation in datapoints collected in the same area at different timepoints (**Supplementary Materials**). We identified influential outliers in the regression model based on a Cook’s distance greater than 4/(total number of studies). In sensitivity analyses, we removed each influential outlier in turn from the model and restricted the dataset to locations with BL data before and after ITN introduction to evaluate the robustness of the findings.

All analyses were conducted in R statistical software (v4.2.2) using the *glmmTMB* (v1.1.7) and *ggeffects* (v1.2.3) packages. MAP estimates of ITN use were extracted from the *foresite* package (v0.1.0, (16)).

## Results

### Study characteristics

The literature search returned a total of 2333 articles and 116 records identified on IARC-associated websites (**Figure 1**). After screening and deduplication of the two sets of data sources, a total of 13 publications from literature databases and 10 IARC reports were included (1, 5, 17-37).

**Figure 1.**
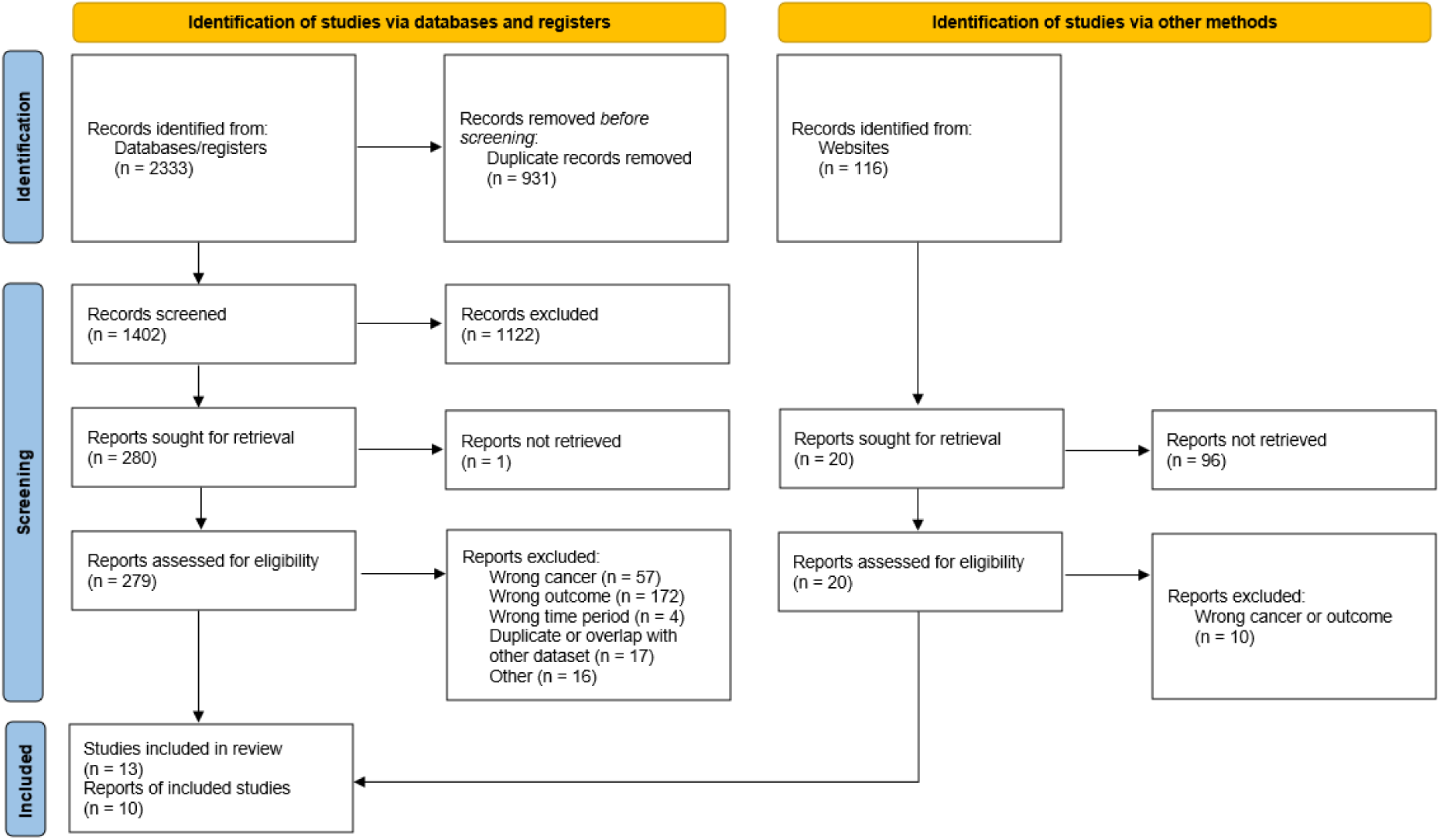
Flow diagram of the systematic review on Burkitt lymphoma incidence rates. Websites associated with the International Agency for Research on Cancer (IARC) included IARC Publications, the Global Cancer Observatory and the African Cancer Registry Network.

76 location- and time-specific BL incidence rates based on a total of 5,700 cases were extracted from the 23 publications. Included data covered the 1990-2017 time period, with slightly more datapoints available after 2005 (**Figure 2A**). 80% of datapoints came from population-based cancer registries. Among the 13 studies identified in the literature search, quality scores ranged from 1 to 4 (highest score; lowest risk of bias). Common limitations included passive case finding and inconsistent or poorly defined confirmation of diagnosis (**Table S2**).

**Figure 2.**
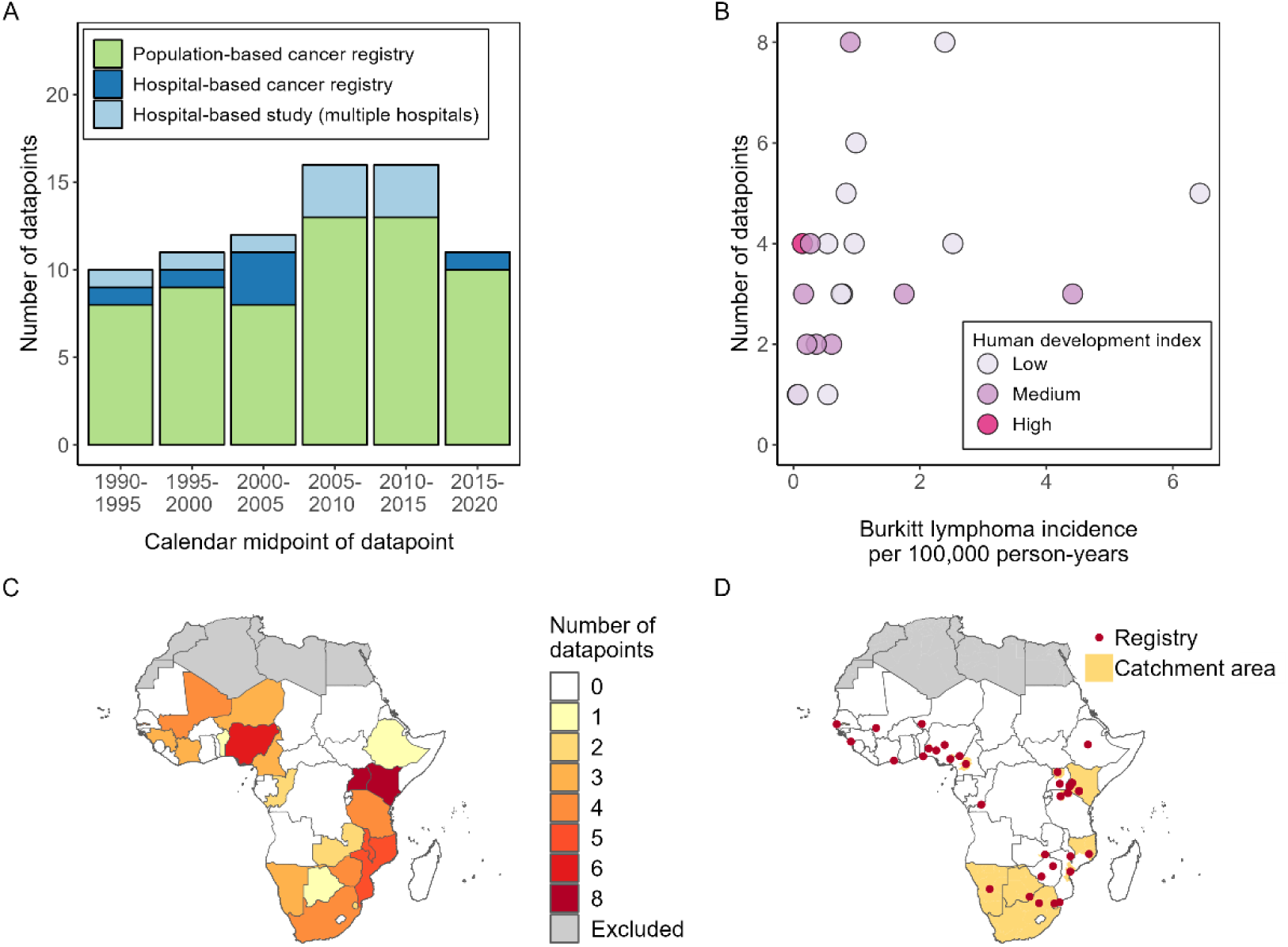
Overview of published Burkitt lymphoma incidence rates between 1990 and 2023. Datapoints apply to different locations and/or different timepoints in a country. (a) Number of datapoints by study type and over time. Cancer data was usually reported for periods of several years, but the number of studies were grouped based on the midpoint of each time period. (b) Number of datapoints by mean reported Burkitt lymphoma incidence rate in each country. Colours represent the human development index level of the country. (c) Number of datapoints in each country. (d) Location and catchment area of the covered population for each incidence datapoint. Yellow shading indicates the first administrative unit covered by the registry or study. Red points indicate the location of the registry or main referral hospital site for each datapoint.

There was a moderate positive correlation between the mean reported BL incidence rate and the number of datapoints in a country, but no relationship between the number of datapoints and the HDI (**Figure 2B**). BL incidence rates came from 32 locations in 21 countries, with 0-8 per country (**Figure 2C**). Many countries in Central and West Africa had no data on BL incidence, while countries in East and Southern Africa had better coverage. The actual catchment area covered in these countries was usually very small (**Figure 2D**). Only The Gambia in West Africa, and South Africa, Eswatini, Botswana and Namibia in Southern Africa had cancer registries with national-level coverage. However, these Southern African countries generally had the lowest BL rates. They were also considered separately, since analyses relating to ITN timing and use were restricted to the 66 other datapoints from locations with large-scale ITN use, covering 17 countries.

### Time trends in BL incidence

The median year of ITN introduction in the included locations was 2005 (range 2004-2011). BL incidence was estimated to be 44% (95% CI 12-64%) lower in the time period after large-scale ITN introduction compared to before. **Table 2** shows pooled incidence rates for each country for the period before (years 1990-2008) and after (years 2007-2017) introduction of ITNs, and across the whole 1990-2017 time period for countries without large-scale ITN use. Across countries, these pooled incidence rates were 1.36 (95% CI 0.88-2.10) and 0.76 (95% CI 0.50-1.16) per 100,000 person years for the periods before and after ITN introduction, respectively. The pooled rate was much lower in countries without ITNs [0.18 (95% CI 0.07-0.46) per 100,000 person-years]. However, there was also large variation in the estimated incidence rates between countries, ranging from 0.09 per 100,000 (95% CI 0.02-0.37) in Mali to 5.98 per 100,000 (95% CI 3.82-9.37) in Malawi before ITN introduction, and from 0.07 per 100,000 (95% CI 0.01-0.43) in Ethiopia to 8.52 per 100,000 (95% CI 2.75-26.33) in Malawi after ITN introduction. 65% of all BL cases in the dataset were recorded in three countries (Kenya, Uganda and Tanzania).

**Table 2.** Country-level pooled estimates of Burkitt lymphoma incidence. Estimates are presented separately for the time periods before and after introduction of insecticide-treated nets (ITNs) and for countries without large-scale use of ITNs. The number of datapoints can refer to different time periods or different studies in the same location. Incidence rates with 95% confidence intervals were calculated using a negative binomial model and adjusted for clustering in data from the same geographical location.

| Country (sources) | Locations | Time periods | Number of datapoints | Number of cases | Person-time at risk | Adjusted incidence rate per 100,000 person-years (95% CI) |
| --- | --- | --- | --- | --- | --- | --- |
| **Time period before introduction of ITNs (1990-2008)** |  |  |  |  |  |  |
| Cameroon (20, 28, 33) | Northwest Province, Yaounde | 2003-2006, 2007-2009, 2004-2006 | 3 | 239 | 6,275,949 | 4.36 (2.57-7.39) |
| Congo (29) | Brazzaville | 1996-1999 | 1 | 11 | 1,017,088 | 1.08 (0.38-3.11) |
| Cote d'Ivoire (29) | Abidjan | 1995-1997 | 1 | 49 | 3,788,100 | 1.29 (0.52-3.24) |
| Guinea (1, 29) | Conakry | 1993-1995, 1996-1999, 2001-2010 | 3 | 52 | 9,029,258 | 0.75 (0.41-1.34) |
| Kenya (1, 23, 26, 29, 33) | Eldoret, Nationwide, Nyanza Province | 1998-2006, 1988-1992, 1993-1997, 1999-2004 | 4 | 1,263 | 134,918,246 | 1.20 (0.77-1.89) |
| Malawi (18, 29, 32, 36) | Blantyre | 1991-1995, 1996-1998, 1999-2001, 2003-2007 | 4 | 403 | 5,472,399 | 5.98 (3.82-9.37) |
| Mali (29) | Bamako | 1988-1997 | 1 | 3 | 3,388,015 | 0.09 (0.02-0.37) |
| Mozambique (21) | Maputo | 1991-2008 | 1 | 133 | 7,130,923 | 1.87 (0.76-4.55) |
| Niger (1, 29, 30) | Niamey | 1993-1999, 2001-2005 | 2 | 29 | 3,366,189 | 0.87 (0.42-1.78) |
| Nigeria (29, 30) | Ibadan | 1993-1999, 2006-2009 | 2 | 68 | 3,534,946 | 1.96 (1.01-3.80) |
| Tanzania (17) | Mwanza and Mara | 2000-2004 | 1 | 540 | 10,114,350 | 5.34 (2.22-12.86) |
| The Gambia (1, 29, 30, 34) | National | 1988-1996, 1997-1998, 2002-2006 | 3 | 45 | 7,965,430 | 0.63 (0.34-1.15) |
| Uganda (25, 27, 34-36) | Northern Uganda, Kyadondo County | 1997-2001, 2002-2006, 1989-1991, | 6 | 836 | 38,319,546 | 2.57 (1.78-3.72) |
|  |  | 1993-1997, 1998-2002, 2003-2007 |  |  |  |  |
| Zambia (19) | Nationwide | 1990-1992 | 1 | 11 | 9,166,667 | 0.12 (0.04-0.34) |
| Zimbabwe (29, 35, 36) | Harare | 1990-1997, 1998-2006 | 2 | 15 | 8,308,590 | 0.18 (0.08-0.40) |
| **Total before introduction of ITNs** |  | **1990-2008** | **35** | **3,697** | **251,795,696** | **1.36 (0.88-2.10)** |
| **Time period after introduction of ITNs (2007-2017)** |  |  |  |  |  |  |
| Benin (31) | Cotonou | 2014-2016 | 1 | 4 | 7,359,00 | 0.54 (0.12-2.39) |
| Congo (30, 31) | Brazzaville | 2009-2016 | 1 | 6 | 4,606,275 | 0.13 (0.03-0.51) |
| Cote d'Ivoire (30, 31) | Abidjan | 2012-2013, 2014-2015 | 2 | 118 | 6,008,792 | 1.98 (0.88-4.43) |
| Ethiopia (1, 31) | Addis Ababa | 2011-2016 | 1 | 3 | 4,207,728 | 0.07 (0.01-0.35) |
| Kenya (1, 5, 30, 31) | Eldoret, Nairobi, Northwestern Kenya | 2007-2011, 2007-2011, 2012-2014, 2012-2016 | 4 | 224 | 27,487,675 | 0.60 (0.33-1.08) |
| Malawi (1, 36) | Blantyre | 2008-2010 | 1 | 102 | 1,197,708 | 8.52 (2.75-26.33) |
| Mali (30, 31, 33) | Bamako | 2005-2009, 2010-2014, 2015-2017 | 3 | 121 | 10,353,780 | 1.25 (0.64-2.43) |
| Mozambique (22, 24, 30, 31) | Beira, Maputo, Northern Region | 2009-2013, 2014-2017, 2014-2017, 2015 | 4 | 35 | 7,730,296 | 0.54 (0.27-1.07) |
| Niger (30) | Niamey | 2006-2009 | 1 | 9 | 1,717,564 | 0.52 (0.14-1.90) |
| Nigeria (30, 31) | Abuja, Calabar, Ekiti | 2013-2016, 2009-2013, 2016-2017, 2013-2017 | 4 | 13 | 4,426,697 | 0.43 (0.19-0.97) |
| Tanzania (5, 17, 31) | Mwanza and Mara | 2005-2009, 2012-2015, 2016-2017 | 3 | 504 | 37,787,627 | 1.63 (0.84-3.16) |
| The Gambia (30) | National | 2007-2011 | 1 | 6 | 3,723,130 | 0.16 (0.04-0.63) |
| Uganda (5, 31, 37) | Kyadondo County, Northern Uganda | 2008-2013, 2010-2016 | 2 | 359 | 18,867,124 | 1.84 (0.83-4.08) |
| Zambia (31) | Lusaka | 2011-2015 | 1 | 13 | 4,246,505 | 0.31 (0.09-1.06) |
| Zimbabwe (1, 31, 36) | Bulawayo, Harare | 2013-2015, 2007-2015 | 2 | 12 | 5,181,909 | 0.30 (0.10-0.87) |
| **Total after introduction of ITNs** |  | **2007-2017** | **31** | **1,529** | **138,278,710** | **0.76 (0.50-1.16)** |
| **Countries without large-scale ITN use (1990-2017)** |  |  |  |  |  |  |
| Botswana (1, 33) | National | 1999-2008 | 1 | 4 | 6,334,397 | 0.06 (0.02-0.19) |
| Eswatini (29, 31) | National | 1989-1999, 2016-2017 | 2 | 12 | 2,484,569 | 0.45 (0.23-0.90) |
| Namibia (29-31) | National | 1995-1998, 2009, 2013-2015 | 3 | 12 | 5,945,760 | 0.20 (0.10-0.38) |
| South Africa (1, 29-31, 33) | National | 2003-2007, 2008-2012, 2013-2016, 1989-1992, 1998-2006, 2007, 2008-2014 | 4 | 446 | 303,370,823 | 0.14 (0.11-0.19) |
| **Total for countries without large-scale ITN use** |  | **1991-2017** | **10** | **474** | **318,135,549** | **0.18 (0.07-0.46)** |

In locations with at least one datapoint before and after ITN introduction, rate ratios were highly variable and time trends were not always consistent across locations (**Figure 3, Figure S2**). The incidence rate appeared to be lower in the period after ITN introduction in 7 out of 12 locations, but confidence intervals overlapped in 3 of these. The largest reductions were seen in Maputo in Mozambique [92% (95% CI 69-98)], Brazzaville in Congo [88% (95% CI 67-96)] and in The Gambia [71% (95% CI 33-88)]. These locations also had among the highest mean ITN use since 2000 (over 75^th^ percentile at 18%, 15% and 19%, respectively). In Bamako in Mali, the incidence rate in the 2005-2017 period was significantly higher than in 1988-1997. In the two registries without large-scale ITN use (Namibia and South Africa), rates were slightly higher after 2005 than before, but there was no significant difference between the two time periods.

**Figure 3.**
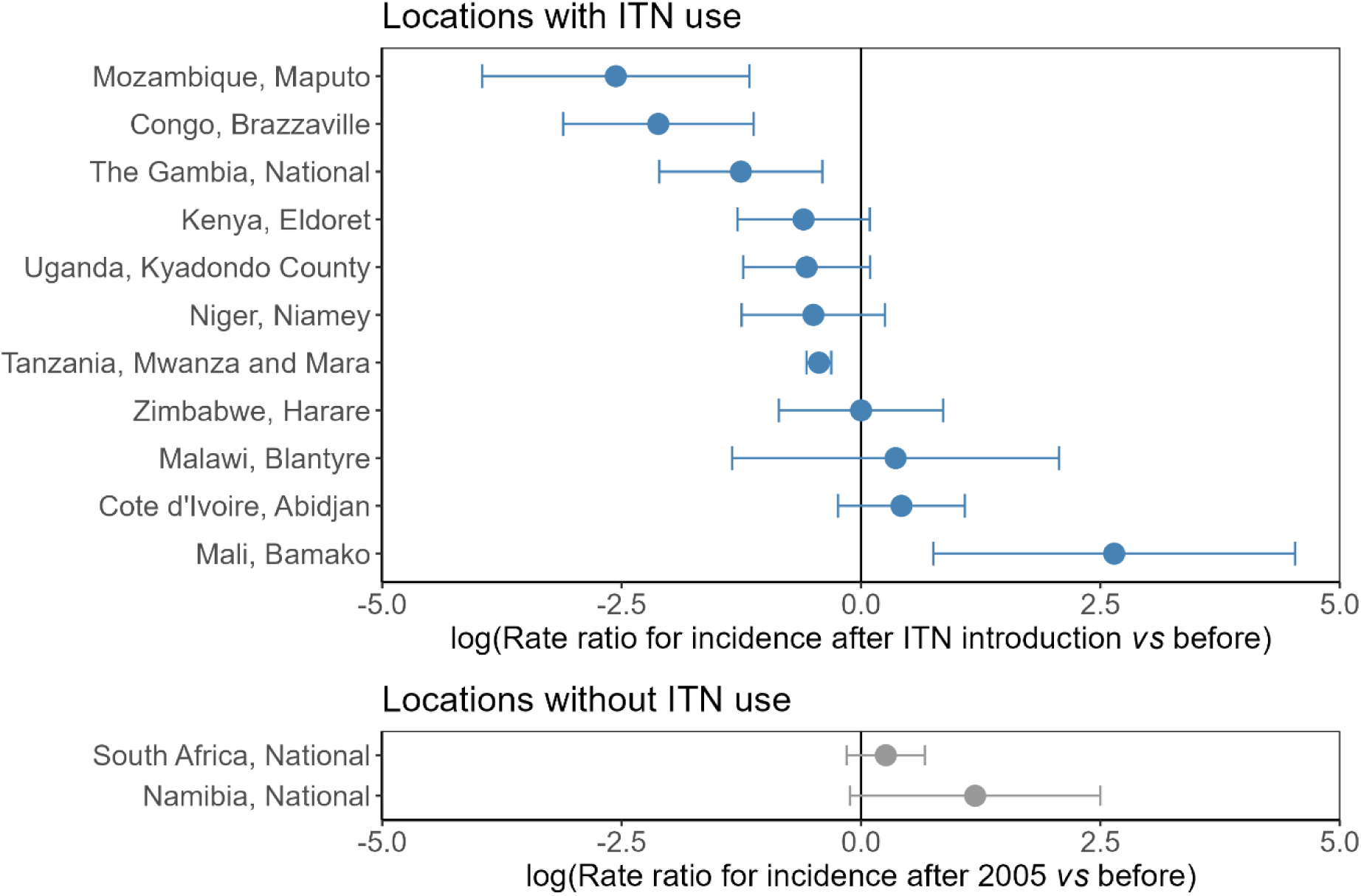
Rate ratios for Burkitt lymphoma incidence after insecticide-treated net (ITN) introduction compared to before in different cancer registries/study locations. Rate ratios are shown on a logarithmic scale for all locations with at least one datapoint in both time periods. Error bars represent the 95% confidence intervals. Namibia and South Africa (grey) did not have wide-scale ITN use but were separated into two time periods before and after 2005 (the median year of ITN introduction).

### Population-level association between ITN use and BL incidence

**Table 3** shows the crude and adjusted associations of BL incidence rates with average population ITN use over 10 years and potential confounders. Univariate and multivariate showed comparable results. There was evidence for a population-level association between ITN use and BL incidence rates after adjusting for baseline malaria prevalence, adult HIV prevalence, country HDI level and population urbanicity (p=0.005). A 1% increase in average ITN use was associated with a 2% (95% CI 1-4%) reduction in BL incidence, as illustrated in **Figure 4**. Baseline malaria prevalence was positively associated with BL incidence after adjusting for all other covariates, but there was no evidence for an association with the other variables (**Table 3**).

**Figure 4.**
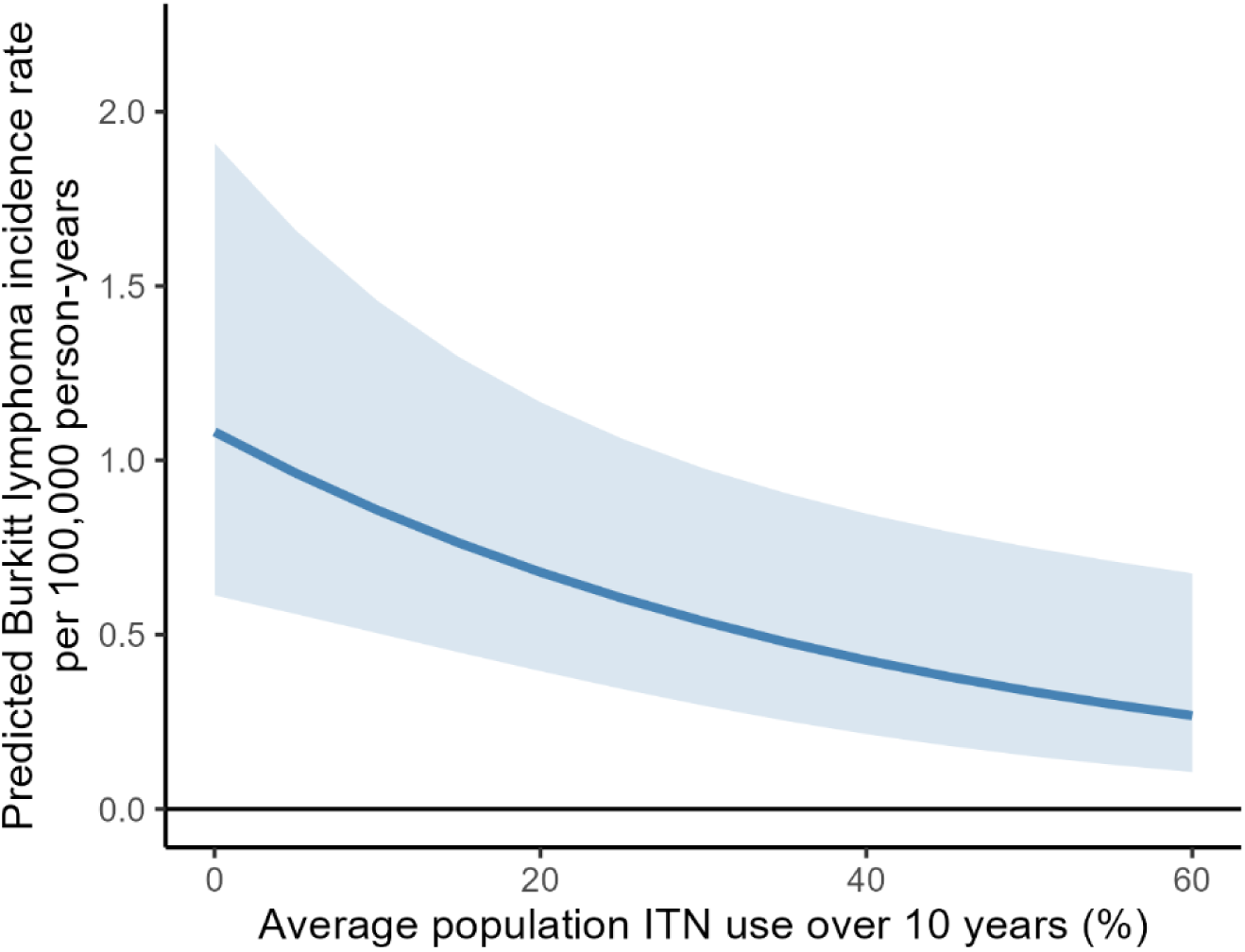
Exposure-response relationship between insecticide-treated net (ITN) use and the Burkitt lymphoma incidence rate in sub-Saharan Africa in the multivariate model. ITN use represents the mean ITN use in the population at the first administrative level in the 10 years before the Burkitt lymphoma data collection period.

**Table 3.** Negative binomial regression models for association between Burkitt lymphoma incidence and insecticide-treated net (ITN) use. ITN use represents the mean ITN use in the population at the first administrative level in the 10 years before the Burkitt lymphoma data collection period. The analysis was conducted on 66 incidence estimates based on 5,226 cases and a total population at risk of 390,074,406.

| **Covariate** | **Crude rate ratio**<br>**(95% CI)** | ***p*-value** | **Adjusted rate ratio**<br>**(95% CI)** | ***p*-value** |
| --- | --- | --- | --- | --- |
| **Average population ITN use over 10 years (%)** | 0.98 (0.96-0.99) | 0.003 | 0.98 (0.96-0.99) | 0.005 |
| **Baseline malaria parasite prevalence in 2000 (%)** | 1.02 (1.00-1.04) | 0.02 | 1.02 (1.01-1.04) | 0.009 |
| **HIV prevalence in 15-49 year olds (%)** | 1.02 (0.96-1.08) | 0.54 | 1.01 (0.96-1.07) | 0.61 |
| **Country Human Development Index level** |  |  |  |  |
| *HDI < 0.45* | 1.00 (reference) |  | 1.00 (reference) |  |
| *HDI ≥ 0.45* | 0.75 (0.43-1.29) | 0.29 | 0.95 (0.56-1.61) | 0.85 |
| **Population urbanicity status** |  |  |  |  |
| *Urban* | 1.00 (reference) |  | 1.00 (reference) |  |
| *Both* | 1.70 (0.77-3.74) | 0.19 | 1.27 (0.62-2.60) | 0.51 |
| *Rural* | 1.92 (0.48-7.72) | 0.36 | 2.17 (0.66-7.16) | 0.20 |

Comparing the main result to different exposure time periods for ITN use, the association was similar for average ITN use in the 5 and 15 years before the BL datapoint [rate ratio 0.98 (95% CI 0.97-0.99) and 0.97 (95% CI 0.95-0.99), respectively]. In line with our hypothesis, concurrent ITN use in the population was not strongly associated with BL incidence (rate ratio 0.99, 95% CI 0.98-1.00) (**Table S3**). In additional sensitivity analyses, removing influential outlier studies and restricting the analysis to locations with BL incidence data before and after ITN introduction did not affect conclusions about the association between ITN use and BL incidence, though the strength of the association was lower in some cases (**Table S4**).

## Discussion

In this study, we investigated changes in BL incidence in sub-Saharan African children in relation to large-scale introduction of ITNs in the early 2000s. In a systematic review, we identified 66 datapoints on BL incidence from locations with large-scale ITN use, comprising 5,226 cancer cases and covering 17 countries between 1990 and 2017. We found BL rates were 44% (95% CI 12-64%) lower in the time period after ITN introduction compared to before. Pooled incidence rates were estimated at 1.36 (95% CI 0.88-2.10) and 0.76 (95% CI 0.50-1.16) per 100,000 person-years before and after ITN introduction, respectively. In an ecological analysis, we described an association between increasing ITN use in the population and decreases in childhood BL incidence after adjusting for potential confounders [rate ratio 0.98 (95% CI 0.96-0.99) for average ITN use in the 10 years before BL data collection].

Previous studies and cancer registries have also noted a decreasing temporal trend in incidence, both for BL (5, 17, 38, 39) and childhood non-Hodgkin lymphoma more generally (39). Given existing evidence for an aetiological role of malaria infection in BL development, the population-level association between ITN use and BL described in this study is plausible. ITNs offer both individual and community-wide protection from malaria infection due to the protective physical barrier and the killing effect of insecticides on mosquitoes, thereby reducing malaria prevalence (40). As previously suggested for malaria infection, our results also point towards past cumulative rather than concurrent exposure to ITNs as a predictor for lower BL incidence on the population level (5, 8).

However, several alternative factors could explain changes in BL incidence over time. Firstly, studies on the effect of antiretroviral treatment (ART) coverage on incidence of HIV-related cancers suggest that HIV trends in children could explain declines in incidence (39, 41). However, concurrent HIV prevalence in adults was not associated with BL incidence in our study, and BL rates can also remain constant in people living with HIV in the ART era (42).

Secondly, the variable time trends seen in individual registries suggest other influences on observed BL incidence, which limit comparability of data from different time periods, locations and sources. The coverage, completeness and diagnostic methods of a registry can vary significantly over time. In Mali, BL incidence rates appeared to be higher in the period after ITN introduction, but the rate before ITN introduction was based on only three cases detected over a 10-year period. Underdiagnosis of childhood cancers was suspected at the time (29), with subsequent improvement in case finding procedures reflected in total cancer numbers (31). Studies of temporal trends in population-based cancer registries in Africa have also reported increases in incidence for other cancer types in adults (43-45). Even though access to diagnosis might generally be expected to improve over time and lead to higher rates, other factors such as intermittent political instability or funding disruption could also contribute to variation in rates (31, 44, 46).

Despite improvements, cancer surveillance in sub-Saharan Africa is still of low quality overall (46). While the historical data used in this study must be interpreted in the context of these limitations, improved cancer surveillance is essential for assessing current and future cancer burden and mortality. Our estimate of BL incidence in the period after ITN introduction can inform planning and targeting of resources in cancer control programmes. It provides further support for the rate reported in a previous study [0.64 per 100,000 person-years in 2018], which extrapolated national estimates of non-Hodgkin lymphoma incidence in countries without primary BL data (47).

Burden estimates are of particular importance because access to effective treatment for childhood cancer faces numerous challenges and remains a low priority for healthcare systems in sub-Saharan Africa (48). Less than a third of children diagnosed with cancer are estimated to survive compared to 80% in high-income countries (49). The possibility that BL incidence may be decreasing in sub-Saharan Africa following successful malaria control programmes therefore has positive implications in terms of a reduced burden on patients and the health system. Given the comparatively higher political commitment and funding for infectious disease programmes like malaria and HIV, a multidisciplinary approach to addressing childhood cancer and infectious diseases has previously been suggested (49, 50). Collaboration with existing malaria programmes could raise awareness, attract funding and increase political commitment to BL prevention, treatment and research (50). Nevertheless, with plateauing funding and rising insecticide and treatment resistance, continued progress in malaria control also needs to remain a priority.

Our study used all publicly available data accrued over almost 30 years and, to the best of our knowledge, provides the first evidence for a population-level decline in childhood Burkitt lymphoma burden in malaria-endemic sub-Saharan Africa associated with large-scale rollout of ITNs for malaria control. These findings provide support for the existing observational evidence base for a potential role of malaria interventions in preventing BL in a context where conducting a randomised controlled trial would be unfeasible and unethical.

Beyond the considerations already outlined, there were several additional limitations. Firstly, BL incidence rates were assembled from various locations, and only a subset of these had data at more than one timepoint. It is not known how representative this published data would be of all incidence rates across malaria-endemic sub-Saharan Africa, and unknown systematic differences between the locations included in the period before ITN introduction compared to after could have biased the results. Secondly, our inclusion criteria were relatively broad to make use of all available data, but different methods in recording of cancer incidence between locations and over time could have affected the results, especially as confirmation of diagnosis was frequently suboptimal (**Table S2**). The small case numbers in some studies also suggest substantial underdiagnosis. Lastly, since an ecological study design was employed, the association between ITN use and BL should not be interpreted to apply on the individual level; both ITN use and potential confounders only represent proxies for the average exposure in the population, and were in many cases based on model estimates.

In conclusion, this study suggests that childhood BL burden in sub-Saharan Africa may be declining as ITN use in the population increases. Analysis of all available data on BL incidence in malaria-endemic African countries indicates that incidence is lower now than before the introduction of mass ITN distribution programmes around 2005. However, data came from various sources and was often of low quality, underlining the need for improved cancer surveillance in sub-Saharan Africa to allow robust investigations of trends in cancer incidence.

## Contributors

NS conceived the study. NS and JK conducted the systematic review and were responsible for data curation. EKA provided domain knowledge for the study and contributed to study design. NS conducted the analysis and wrote the first draft of the manuscript. All authors contributed to interpreting the results, reviewing and revising the manuscript, and approved the final version for submission.

## Data availability statement

All data reported in this study were extracted from published studies. All covariate estimates used in the analysis are publicly available from the sources cited in the article.

