## Supplementary Material for "Association between insecticide-treated bed net use and Burkitt lymphoma incidence in children in sub-Saharan Africa: a systematic review and population-level analysis"

### List of tables and figures

**Figure S1.** Estimated insecticide-treated net (ITN) use between 2000 and 2020 for each location with data on Burkitt lymphoma. .......... 7
**Figure S2**. Burkitt lymphoma incidence rate in the time period before and after introduction of ITNs for locations with at least one datapoint in each period .......... 12

### Supplementary Methods

#### Systematic review

##### Search strategy

**Table S1** shows the search terms for the systematic review conducted in the EMBASE, Medline and Global Health databases. We also searched for publications on websites associated with the International Agency for Research on Cancer (1-3).

**Table S1.** Search strategy for EMBASE, Medline and Global Health databases. The search strategy was developed and refined after cross-referencing of a pilot search against a list of known relevant papers. The original study protocol registered in PROSPERO (reference CRD42023411961) also included the African Journals Online database, but due to limitations in applying reproducible search strategies it was excluded.

| | |
|---|---|
| **Database: EMBASE** | |
| 1 | "Africa south of the Sahara"/ |
| 2 | (africa* or SSA or angola* or benin* or botswana* or burkina faso* or burundi* or cabo verd* or cameroon* or cameroun or cape verd* or central african republic* or chad* or comoros or (congo* not congo red) or cote divoir* or democratic republic of the congo* or djibouti* or equatorial guinea* or eritrea* or ethiopia* or gabon* or gambia* or ghan* or (guinea* not guinea pig* not guinea worm*) or guinea-bissau* or ivory coast or kenya* or lesotho* or liberia* or madagascar* or malawi* or mali or maurit* or mauritania* or mozambi* or namibia* or niger* or nigeria* or reunion or rwanda* or (sao tome and principe*) or senegal* or seychelle* or sierra leone* or somali* or south africa* or south sudan* or (sudan* not sudan blue) or swazi* or eswatini or tanzania* or togo* or uganda* or zambia* or zimbabwe*).mp. [mp=title, abstract, heading word, drug trade name, original title, device manufacturer, drug manufacturer, device trade name, keyword heading word, floating subheading word, candidate term word] |
| 3 | 1 or 2 |
| 4 | Burkitt lymphoma/ |
| 5 | ((burkitt* adj3 lymphoma*) or burkitt* lymphoma* or african lymphoma* or african malignant lymphoma* or burkitt* disease or burkitt* like lymphoma* or burkitt* tumor* or burkitt* tumour* or Lymphoma*, African or Lymphoma*, burkitt* or Malignant lymphoma*, African or non-hodgkin* lymphoma*).mp. [mp=title, abstract, heading word, drug trade name, original title, device manufacturer, drug manufacturer, device trade name, keyword heading word, floating subheading word, candidate term word] |
| 6 | 4 or 5 |
| 7 | (incidence or rate* or incident or distribution or pattern or prevalence or distribution or patterns or frequency).mp [mp=title, abstract, heading word, drug trade name, original title, device manufacturer, drug manufacturer, device trade name, keyword heading word, floating subheading word, candidate term word] |
| 8 | 3 and 6 and 7 |
| 9 | limit 8 to yr="1990 -Current" |
| **Database: Medline** | |
| 1 | "Africa South of the Sahara"/ |
| 2 | (africa* or SSA or angola* or benin* or botswana* or burkina faso* or burundi* or cabo verd* or cameroon* or cameroun or cape verd* or central african republic* or chad* or comoros or (congo* not congo red) or cote divoir* or democratic republic of the congo* or djibouti* or equatorial guinea* |

| | |
|---|---|
| | or eritrea* or ethiopia* or gabon* or gambia* or ghan* or (guinea* not guinea pig* not guinea worm*) or guinea-bissau* or ivory coast or kenya* or lesotho* or liberia* or madagascar* or malawi* or mali or maurit* or mauritania* or mozambi* or namibia* or niger* or nigeria* or reunion or rwanda* or (sao tome and principe*) or senegal* or seychelle* or sierra leone* or somali* or south africa* or south sudan* or (sudan* not sudan blue) or swazi* or eswatini or tanzania* or togo* or uganda* or zambia* or zimbabwe*).mp. |
| 3 | 1 or 2 |
| 4 | Burkitt lymphoma/ |
| 5 | ((burkitt* adj3 lymphoma*) or burkitt* lymphoma* or african lymphoma* or african malignant lymphoma* or burkitt* disease or burkitt* like lymphoma* or burkitt* tumor* or burkitt* tumour* or Lymphoma*, African or Lymphoma*, burkitt* or Malignant lymphoma*, African or non-hodgkin* lymphoma*).mp. |
| 6 | 4 or 5 |
| 7 | (incidence or rate* or incident or distribution or pattern or prevalence or distribution or patterns or frequency).mp. [mp=title, book title, abstract, original title, name of substance word, subject heading word, floating sub-heading word, keyword heading word, organism supplementary concept word, protocol supplementary concept word, rare disease supplementary concept word, unique identifier, synonyms, population supplementary concept word, anatomy supplementary concept word] |
| 8 | 3 and 6 and 7 |
| 9 | limit 8 to yr="1990 -Current" |
| **Database: Global Health** | |
| 1 | "Africa South of the Sahara"/ |
| 2 | (africa* or SSA or angola* or benin* or botswana* or burkina faso* or burundi* or cabo verd* or cameroon* or cameroun or cape verd* or central african republic* or chad* or comoros or (congo* not congo red) or cote divoir* or democratic republic of the congo* or djibouti* or equatorial guinea* or eritrea* or ethiopia* or gabon* or gambia* or ghan* or (guinea* not guinea pig* not guinea worm*) or guinea-bissau* or ivory coast or kenya* or lesotho* or liberia* or madagascar* or malawi* or mali or maurit* or mauritania* or mozambi* or namibia* or niger* or nigeria* or reunion or rwanda* or (sao tome and principe*) or senegal* or seychelle* or sierra leone* or somali* or south africa* or south sudan* or (sudan* not sudan blue) or swazi* or eswatini or tanzania* or togo* or uganda* or zambia* or zimbabwe*).mp. [mp=abstract, title, original title, heading words, cabicodes words] |
| 3 | 1 or 2 |
| 4 | Burkitt's lymphoma.sh. |
| 5 | ((burkitt* adj3 lymphoma*) or burkitt* lymphoma* or african lymphoma* or african malignant lymphoma* or burkitt* disease or burkitt* like lymphoma* or burkitt* tumor* or burkitt* tumour* or Lymphoma*, African or Lymphoma*, burkitt* or Malignant lymphoma*, African or non-hodgkin* lymphoma*).mp. [mp=abstract, title, original title, heading words, cabicodes words] |
| 6 | 4 or 5 |
| 7 | (incidence or rate* or incident or distribution or pattern or prevalence or distribution or patterns or frequency).mp. [mp=abstract, title, original title, heading words, cabicodes words] |
| 8 | 3 and 6 and 7 |
| 9 | limit 8 to yr="1990 -Current" |

##### Inclusion and exclusion criteria

Epidemiological studies reporting the incidence rate of Burkitt lymphoma in children aged 0-15 years in malaria-endemic sub-Saharan African countries were included. We applied no restrictions on study design, but modelling studies, such as those from the Global Burden of Disease Study, were excluded. To obtain incidence rates among the general population in sub-Saharan Africa, we excluded populations selected for having specific known risk factors for Burkitt lymphoma (e.g. HIV-positive patients) or incidence rates calculated only among hospital patients. Populations residing outside of sub-Saharan Africa, e.g. immigrants of African origin, were also excluded.

Data on cancer cases was included if it was collected prospectively for the purpose of the study or extracted from medical records, and if it referred to a defined location and time period. Retrospective studies assessing cancer burden exclusively based on questionnaires administered to physicians were excluded.

The outcome of interest were incident cancer cases diagnosed as Burkitt lymphoma during a time period with a midpoint in or after 1990. Articles reporting only the frequency of Burkitt lymphoma cases without a defined catchment population at risk were excluded.

**Data processing**

Where necessary, missing quantities (e.g. person-time at risk or incidence rate) were derived from the available information in the articles. Studies identified from the published literature were cross-checked against the dataset extracted from cancer registry publications for duplicates and overlap in data from the same registry. If case counts were sufficiently large (>10 in each period), we stratified exactly overlapping data from the same registry but two different sources into two separate shorter time periods. Data from two partially overlapping time periods were combined by calculating the mean incidence for the longer combined time period.

**Quality assessment**

For studies identified from the literature, quality was assessed based on three predefined criteria: the methods for data collection or cancer registration procedures, case ascertainment and diagnostic methods (including coding criteria), and calculation of person-time at risk. A score of 0 or 1 was assigned for each of these, and stated strengths and limitations were also considered. The total score was used to identify any particularly low-quality studies (score < 1). Quality assessment was not conducted on data extracted from IARC publications, as these all came from population-based cancer registries and had undergone a rigorous editorial process including quality control evaluation (4, 5). However, information on quality considerations for each registry was reviewed for notable deviations.

#### Statistical analysis

**Regression model for the association between ITN use and BL incidence**

We assumed that the observed number of cancer cases *y* follows a negative binomial distribution with mean $\mu$ and dispersion parameter $\theta$, and accounted for clustering of datapoints collected in the same geographical location at different timepoints and for potential population-level confounders. The model is represented by the following set of equations:

$$y \sim NegBin(\mu, \theta)$$

$$\mu = exp(\beta_0 + \beta_1(ITN\ use) + \beta_2(PfPR2000) + \beta_3(HIV\ prevalence) + \beta_4(HDI\ level) + \beta_5(urbanicity) + \ln\text{(pop)} + u)$$

$$u \sim N(0, \sigma_u^2)$$

$$\sigma^2 = \mu(1 + \frac{\mu}{\theta})$$

Where $\beta_0$ is the intercept, $\beta$ parameters are the regression coefficients for the given fixed-effect predictor variables, *ln(pop)* is an offset to account for person-time at risk, and $u$ is the location-specific random effect. $\sigma^2$denotes the variance of the negative binomial distribution.

**Software**

The systematic review was conducted on the Covidence platform (6). All analyses were conducted in R (v4.2.2, (7)). ITN use estimates were extracted from the *foresite* package (v0.1.0, (8)). The *glmmTMB* (v1.1.7) and *ggeffects* packages (v1.2.3) were used for the statistical analysis (9, 10).

### Supplementary Results

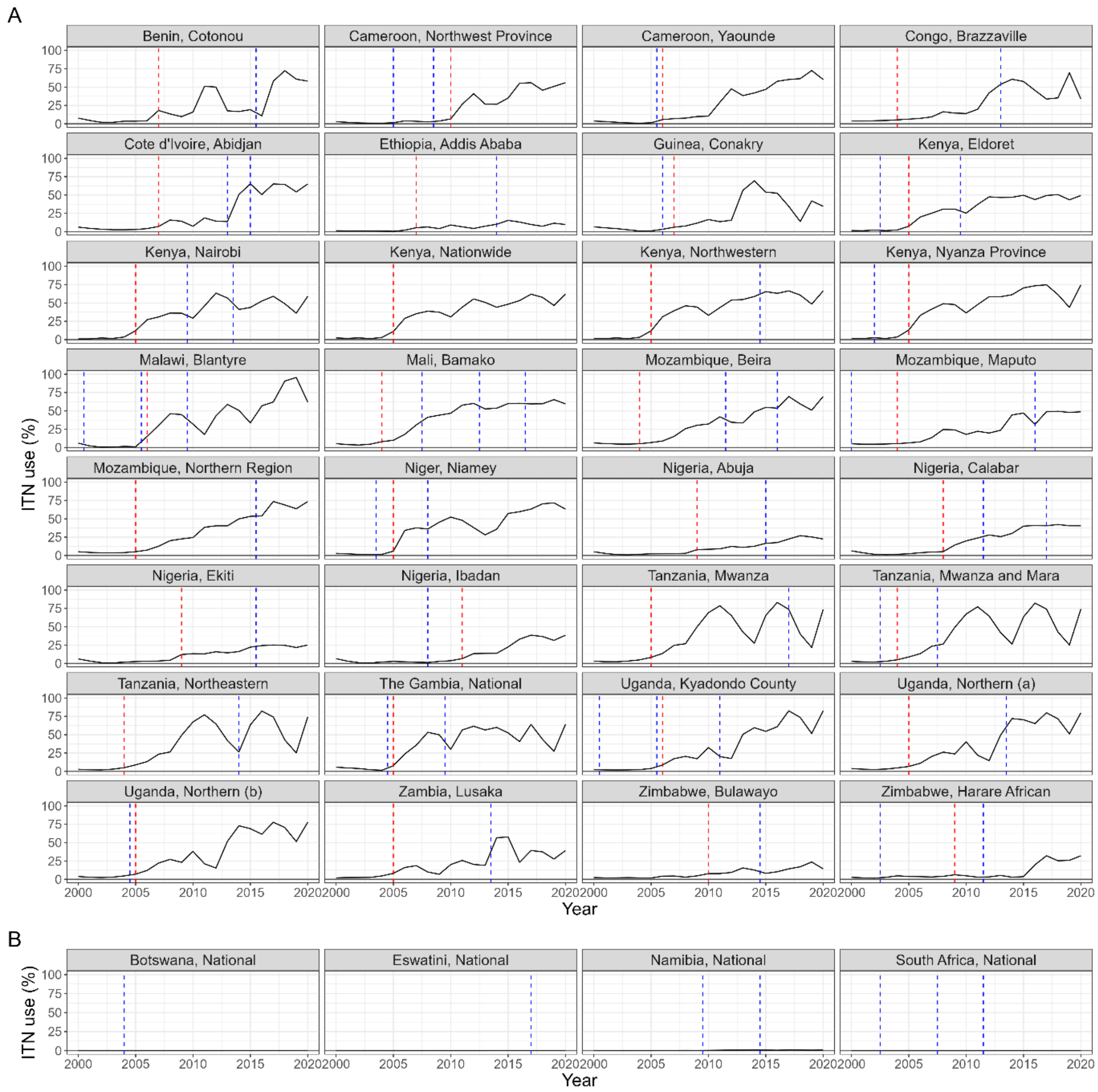

**Figure S1.** Estimated insecticide-treated net (ITN) use between 2000 and 2020 for each location with data on Burkitt lymphoma. (A) Locations with large-scale population-wide ITN use. (B) Locations in southern Africa without large-scale ITN use (<1% in all years). Blue vertical lines represent the calendar midpoint for Burkitt lymphoma rates after 2000. Red vertical lines indicate the timepoint considered as the year of ITN introduction in the analysis.

**Table S2**. Quality assessment of included studies from the literature. The total score is the sum of individual scores in each column, with a possible range from -1 (highest risk of bias) to 4 (lowest risk of bias). BL = Burkitt lymphoma.

| Study | Type of study | Data collection [score] | Confirmation of diagnosis [score] | Calculation of person-time at risk [score] | Notable stated strengths and limitations [score] | Score |
|---|---|---|---|---|---|---|
| Aka et al., 2012, Pediatric Blood & Cancer | Hospital-based study (multiple hospitals) | Case information compiled by a research assistant from routine hospital records from 6 hospitals in Mara and Mwanza regions. The hospitals included were all those able to diagnose and treat BL in the regions. Only cases with a home address in the Mara and Mwanza regions were included. Cases from these two regions that may have been treated at a hospital outside the region were not searched for. [1] | Predominantly clinical diagnosis with confirmation using fine needle aspiration in a few cases. [0] | Annual age-specific population size was extrapolated forward and backwards from the 2002 national census according to region-specific growth rates. [1] | NA [0] | 2 |
| Wright et al., 2009, Tropical Doctor | Hospital-based study (multiple hospitals) | Case data collected from all 16 hospitals, private clinics, the regional pathologist and the Delegation of Public Health in the Northwest province. Only cases in 2015 were included as it was the only year where all hospitals provided data on BL. [1] | Cases were included if they had either a histological diagnosis (70%) or a clinical diagnosis of a fast-growing tumour, that responded to the Malawi 2002 protocol with complete regression within six chemotherapy doses, or, if they died before, an initial response of >50% reduction within three doses. [1] | Population data provided by the Delegation of Public Health, no further information. [1] | NA [0] | 3 |
| Ogwang et al., 2008, International Journal of Cancer | Hospital-based cancer registry | Case data collected in hospital-based cancer registry of the only referral hospital in the region with facilities to both diagnose and treat BL. Only cases with an address in the 10 neighbouring districts were included, for a time period where registry data was considered reasonably complete. [0] | Cases are diagnosed clinically and confirmed using cytology or histology by a senior pathologist at Makerere University Medical School in Kampala. [1] | Annual age-specific population projections were obtained from the Uganda Bureau of Statistics, which were based on population counts from the 1991 and 2002 censuses. [1] | Civil unrest in 1997-2001. Data patterns suggest underreporting (e.g. lower incidence in counties further from Lacor especially during period of civil unrest). [-1] | 1 |
| Broen et al., 2023, Proceedings of the National | Hospital-based study (multiple hospitals, population- | BL cases were recruited at 6 local district or regional hospitals serving a population living in defined geographic areas (2 regions in each country). Only cases who were usual residents (≥4 months prior to | Cases were defined histologically or cytologically (61%), and when this was not possible, according to clinical features, imaging and laboratory | Age-specific population data obtained from each country's statistical bureau and interpolated using district or regional average population | Fulltime field staff were hired to implement the study in the three countries and trained by the | 3 |

| | | | | | | |
|---|---|---|---|---|---|---|
| Academy of Sciences of the United States of America | based case control study) | enrollment) of the study area were included. To increase ascertainment and encourage referral of suspected cases to the six participating hospitals, health education messages about BL were developed and disseminated in the study area. [1] | results compatible with a diagnosis of BL. [1] | growth rates. National census data was available from 2009 in Kenya and 2002 and 2014 in Uganda. [1] | same instructors at Makerere University College of Health Sciences in Uganda. [0] | |
| Lorenzoni et al., 2020, Internationa l Journal of Cancer | Population-based cancer registry | Case data collected from 3 hospitals (including the national referral hospital responsible for most cancer cases diagnoses and registrations), the Private Laboratory of Pathology and death certificates. Death registration is mandatory and death certificates are completed by a physician (for hospital deaths) or the statistical office in the mortuary (for deaths at home). For deaths at home, cause of death information is based on documents brought by the family, and questions to them similar to verbal autopsy. Deaths from cancer are matched with the registry database and hospital records. Only cases resident in Maputo City were included. The Canreg5 system for deduplication, data management and analysis was used. [1] | Tumour site and histology coded according to ICD-O-3 and converted to ICD-10 for analysis. 74% of all non-Hodgin lymphoma diagnoses were morphologically verified and 24% were based on death certificates only. [1] | Annual age-specific population size was interpolated based on the the census of 2007 and 2017 using a constant age- and sex-specific growth rate. [1] | NA [0] | 3 |
| O'Callaghan -Gordo et al., 2016, The American Journal of Tropical Medicine and Hygiene | Hospital-based cancer registry | Prospective collection of all new BL diagnoses in Nampula Central Hospital, which is the referral hospital and has the only pathology department in the region. Only cases living in the region were included. [0] | Diagnosis was confirmed using fine-needle aspiration cytology (82%) or histology (18%) in all but 2 cases. Diagnosis was confirmed by two pathologists. [1] | Age-specific population estimates were obtained from the official projections of the National Institute of Statistics of Mozambique, based on data from the censuses conducted in 1997 and 2007. [0] | NA [0] | 1 |
| Lewis et al., 2012, Paediatrics and | Hospital-based study (multiple hospitals) | Case data collected in the three treatment centers for BL in the country by a specialist BL nurse and a standardised inpatient work-up. Cases | Diagnosis confirmed by fine needle aspiration or based on strict clinical criteria pertaining to a fast growing | Age-specific population size in the 2007 census obtained from the Delegation of Public Health | In Northwest Province, the two BL treatment centres have a long-standing | 4 |

| | | | | | | |
|---|---|---|---|---|---|---|
| International Child Health | | were classified according to their home district. [1] | tumour responding to BL treatment. [1] | and extrapolated using a constant growth rate. [1] | history, a well established education programme for clinicians and patients and provide treatment free of charge. This suggests referral from local health centers is likely. [1] | |
| Rainey et al., 2007, International Journal of Cancer | Hospital-based cancer registry | Case data collected from medical records of all BL cases admitted to Nyanza Provincial General Hospital (the referral center for childhood cancer cases and only treatment center for BL in the region) with a physician diagnosis of Burkitt's lymphoma. Only cases residing in Nyanza Province were included and duplicate cancer records were excluded. [0] | All diagnoses histologically confirmed. For cases with incomplete histology records, inclusion was restricted to those treated for BL and responding to treatment with no competing diagnosis. [1] | Age-specific population estimates based on 1999 census data obtained from the Kenya Medical Research Institute/Wellcome Trust Collaborative Programme and 2000–2004 age-specific population projections generated by the Kenyan Central Bureau of Statistics. [1] | Cancer cases from isolated communities were captured in the hospital records, suggesting that the majority of cases in the region were detected. [0] | 2 |
| Banda et al., 2001, Tropical Medicine & International Health | Population-based cancer registry | Case data collected using active and passive methods of case-finding, including: reports on all cancer cases diagnosed in the central pathology laboratory at Queen Elizabeth Central Hospital (the only one in the country), visits to inpatient wards and oncology outpatient clinics of the hospital and seven other hospitals in the district (including private hospitals), and searching of the register of cancer patients who die in Queen Elizabeth Central Hospital. Only cases resident in Blantyre Districts for at least 6 months were included. The Canreg system for deduplication, data management and analysis was used. [1] | Tumour site and histology coded according to ICD-O-2 and converted to ICD-10 for analysis. 77% of all childhood cancer cases were morphologically verified. [1] | Age-specific population size was interpolated between census data from 1987 and 1998. [1] | NA [0] | 3 |
| Chintu et al., 1995, Archives of | Hospital-based cancer registry | Case data collected from all histopathological records at the University Teaching Hospital, which is | All diagnoses were based on histopathology. [1] | Age-specific population size in the whole country was obtained | NA [0] | 1 |

| | | | | | | |
|---|---|---|---|---|---|---|
| Disease in Childhood | | estimated to cover 77% of the total paediatric population of Zambia. Duplicates and cases with "doubtful information" on diagnosis were excluded. [0] | | from projections based on the 1980 and 1990 census. [0] | | |
| Wabinga et al., 1993, International Journal of Cancer | Population-based cancer registry | Case data collected from histopathological records from the only pathology service in Uganda with diagnostic histological, cytological and autopsy capacity, and active case finding through monthly visits by the cancer registrar to the 4 major hospitals in Uganda to which cancer cases might be admitted (Mulago, Mengo, Rubaga and Nsambya). Only cases resident in Kyadondo County for at least 1 year were included. [1] | Tumour site and histology coded according to ICD-O and converted to ICD-9 for analysis. 71% of all lymphoma cases were confirmed histologically/cytologically and 26% were based solely on clinical examination. [1[ | Age-specific population size assumed to be the same in each year as in the 1991 census. [1] | NA [0] | 3 |
| Lorenzoni et al., 2015, PLoS One | Hospital-based cancer registry | Case data collected from all cancer cases registered in the Department of Pathology of the Maputo Central Hospital, which is the national referral hospital. Cases from sites other than Maputo and duplicate registrations were excluded. [0] | Tumour site and histology coded according to ICD-O and converted to ICD-10 for analysis. All diagnoses were based on histopathology. [1] | Age-specific population size was interpolated between national censuses in 1980, 1997 and 2007 assuming a constant sex- and age-specific growth rate. [1] | NA [0] | 2 |
| Mwanda et al., 2004, East African Medical Journal | Hospital-based study (multiple hospitals) | Case data collected from seven provincial hospitals (Coast General Hospital, Garissa, Embu, Nyeri, Nakuru, Kakamega, Kisumu) and the national referral hospital with BL treatment facilities (Kenyatta National Hospital). Medical records were reviewed between 1988-1992 and data was collected prospectively between 1993-1997. Duplicate registrations were excluded and residence was assigned based on the previous 3 months. [1] | Diagnosis was primarily histological using fine needle aspiration and corroborated by clincial presentation and treatment results. [1] | Age-specific population size estimated based on census data provided by Kenya National Population Census Offices (no further information). [1] | Difficulty in ascertaining residence, particularly in Nairobi, and questions regarding denominator quality. Better case ascertainment in the prospective than the retrospective period. [-1] | 2 |

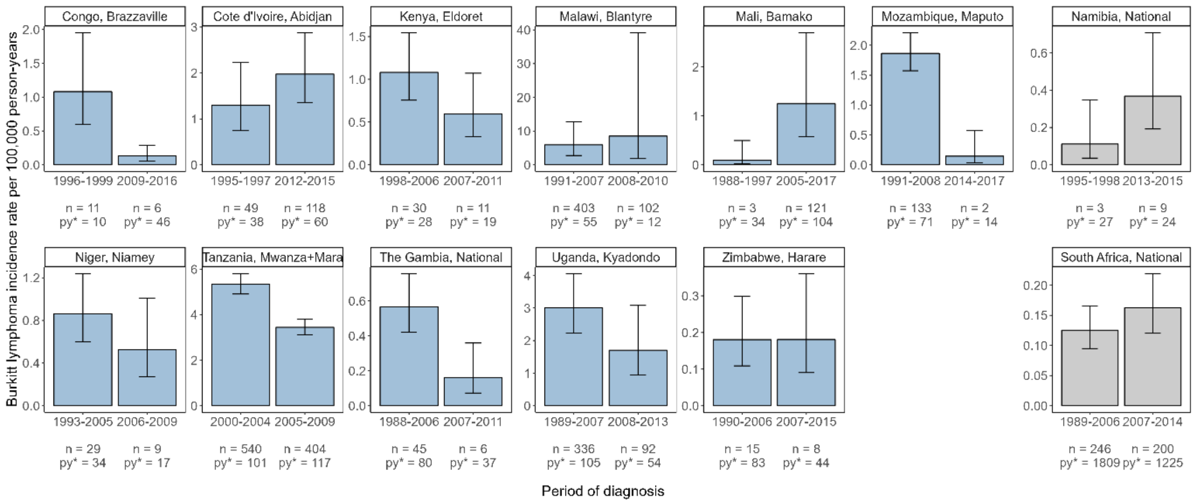

**Figure S2**. Burkitt lymphoma incidence rate in the time period before and after introduction of ITNs for locations with at least one datapoint in each period. Namibia and South Africa (grey bars) did not have wide-scale insecticide-treated net use but were separated into two time periods with midpoints before and after 2005. Incidence rates with 95% confidence intervals were calculated using a negative binomial model and adjusted for clustering in data from the same geographical location. The number of cases (n) and person-years at risk in 100,000s (py*) are given for each time period. The indicated period of diagnosis represents the minimum and maximum year covered in the dataset but may not be fully covered by data.

**Table S3**. Sensitivity analysis for association between Burkitt lymphoma incidence and insecticide-treated net (ITN) use with different ITN use exposure periods.

| ITN use predictor | Average population ITN use over 5 years | | Average population ITN use over 15 years | | Concurrent ITN use | |
|---|---|---|---|---|---|---|
| **Covariate** | **Adjusted rate ratio (95% CI)** | **p-value** | **Adjusted rate ratio (95% CI)** | **p-value** | **Adjusted rate ratio (95% CI)** | **p-value** |
| **ITN use predictor (%)** | 0.98 (0.97-0.99) | 0.005 | 0.97 (0.95-0.99) | 0.004 | 0.99 (0.98-1.00) | 0.09 |
| **Baseline malaria parasite prevalence in 2000 (%)** | 1.02 (1.01-1.04) | 0.01 | 1.02 (1.01-1.04) | 0.008 | 1.03 (1.01-1.04) | 0.008 |
| **HIV prevalence in 15-49 year olds (%)** | 1.01 (0.96-1.07) | 0.67 | 1.01 (0.96-1.07) | 0.60 | 1.02 (0.97-1.08) | 0.47 |
| **Country Human Development Index level** | | | | | | |
| *HDI < 0.45* | 1.00 (reference) | | 1.00 (reference) | | 1.00 (reference) | |
| *HDI ≥ 0.45* | 0.97 (0.57-1.65) | 0.91 | 0.95 (0.56-1.61) | 0.85 | 0.92 (0.53-1.57) | 0.75 |
| **Population urbanicity status** | | | | | | |
| *Urban* | 1.00 (reference) | | 1.00 (reference) | | 1.00 (reference) | |
| *Both* | 1.26 (0.61-2.58) | 0.53 | 1.26 (0.62-2.58) | 0.52 | 1.26 (0.62-2.56) | 0.53 |
| *Rural* | 2.11 (0.64-7.00) | 0.22 | 2.17 (0.66-7.13) | 0.20 | 2.30 (0.71-7.44) | 0.17 |

**Table S4**. Sensitivity analysis for association between Burkitt lymphoma incidence and insecticide-treated net (ITN) use. n indicates the number of incidence datapoints included in each model. Models 1, 2 and 3 included 4282, 5226 and 2473 cancer cases, respectively. The influential outliers, based on a Cook's distance of greater than 4/(total number of studies), were Burkitt lymphoma incidence in Mara and Mwanza, Tanzania, 2000-2009 (11) and Abuja population-based cancer registry, Nigeria, 2013-2016 (4). The study set in Tanzania contributed 17% of all cancer cases in the dataset and, while not among the lowest-scoring studies for quality overall (Table S3), was the only one where diagnosis was almost exclusively based on clinical assessment. Abuja cancer registry recorded zero Burkitt lymphoma cases, and quality concerns about the data from this registry were raised in the publication.

| | **Model 1: Remove influential outlier 1 (n = 64)** | | **Model 2: Remove influential outlier 2 (n = 65)** | | **Model 3: Only locations with BL data before and after ITN introduction (n = 35)** | |
|---|---|---|---|---|---|---|
| **Covariate** | **Adjusted rate ratio (95% CI)** | **p-value** | **Adjusted rate ratio (95% CI)** | **p-value** | **Adjusted rate ratio (95% CI)** | **p-value** |
| **Average population ITN use over 10 years (%)** | 0.98 (0.97- 1.00) | 0.05 | 0.98 (0.96-0.99) | 0.002 | 0.97 (0.95-1.00) | 0.05 |
| **Baseline malaria parasite prevalence in 2000 (%)** | 1.03 (1.01- 1.05) | 0.002 | 1.03 (1.01-1.05) | 0.0009 | 1.04 (1.01-1.07) | 0.01 |
| **HIV prevalence in 15-49 year olds (%)** | 1.02 (0.97- 1.08) | 0.40 | 1.01 (0.96-1.06) | 0.59 | 0.98 (0.93-1.04) | 0.54 |
| **Country Human Development Index level** | | | | | | |
| *HDI < 0.45* | 1.00 (reference) | | 1.00 (reference) | | 1.00 (reference) | |
| *HDI ≥ 0.45* | 1.05 (0.61-1.81) | 0.86 | 1.05 (0.63-1.73) | 0.86 | 0.82 (0.44-1.56) | 0.55 |
| **Population urbanicity status** | | | | | | |
| *Urban* | 1.00 (reference) | | 1.00 (reference) | | 1.00 (reference) | |
| *Both* | 1.05 (0.52- 2.10) | 0.90 | 1.07 (0.53-2.14) | 0.86 | 3.36 (1.37-8.24) | 0.008 |
| *Rural* | 2.41 (0.79-7.38) | 0.12 | 2.09 (0.67-6.58) | 0.21 | 0.89 (0.20-4.00) | 0.88 |
